# A cross-sectional study of digital mental health literacy and communication, predictors of the TAM, and willingness to use smartphone mental health applications among adults in Bangladesh

**DOI:** 10.64898/2026.09.21.26363570

**Authors:** Shabbir Abdullah Maruf, MD Habibur Rahman, Mohammad Biyazid Khan, Md. Al Mamunur Rashid, Tasfia Afrin Faria, Hossain Mahmud, Most Sakila Azmin

## Abstract

**Background:** Digital mental health applications offer scalable pathways to expand mental healthcare access in low- and middle-income countries, yet limited evidence exists on factors predicting willingness to use such applications in South Asia. This study examined associations between digital mental health literacy (DMHL), Technology Acceptance Model (TAM) constructs, mental health symptoms, and willingness to use smartphone-based mental health applications among adults in Bangladesh.

**Methods:** Cross-sectional study conducted from February to June 2026 across urban (Dhaka), semi-urban (Chattogram), and rural (Sylhet) divisions of Bangladesh. Adults aged 18–60 years attending primary care facilities with smartphone access (n = 400) completed a face-to-face interview in Bangla assessing sociodemographic, depression (PHQ-9), anxiety (GAD-7), DMHL (6-item scale, α = .762), TAM perceived usefulness (5-item, α = .937) and ease of use (5-item, α = .924), and willingness to use mental health applications (5-item scale, α = .946). Hierarchical binary logistic regression examined associations with willingness (dichotomized at mean ≥3.5).

**Results:** Greater DMHL was associated with higher willingness (adjusted odds ratio [aOR] = 1.24, 95% CI: 1.11– 1.38). Both perceived usefulness (aOR = 1.29, 95% CI: 1.18–1.41) and perceived ease of use (aOR = 1.17, 95% CI: 1.07–1.28) independently predicted willingness. Anxiety symptoms were positively associated (aOR = 1.08 per 1-point increase, 95% CI: 1.01–1.15), whereas depression symptoms, age, and sex were not independently significant after adjustment for other variables.

**Conclusions:** Digital literacy, perceived utility of technology, and symptom-specific phenomenology but not demographics alone are associated with willingness to use digital mental health applications. Building DMHL, optimizing the usefulness and usability of the app, and overcoming implementation barriers should all be given priority.

## 1. Introduction

Depression and anxiety have been recognized as major drivers of disability worldwide and have significant disease burden in low- and middle-income countries (LMICs) where service provision is most limited (Kim et al., 2023). The prevalence of mental health problems ranges from 6.5% to 31% among adults in Bangladesh, with around 16% of the population suffering from common mental disorders (CMDs), mainly depression and anxiety disorders (Alam et al., 2025; Sifat et al., 2022). Despite this high prevalence, there is still a severe lack of mental health care in terms of inadequate professional services, lack of mental health literacy, stigma, and a lack of timely access to evidence-based care (Alam et al., 2025; Koly et al., 2022; Sifat et al., 2022).

In recent years, with the advancement of digital technology, large-scale mental health service needs have been met in resource-constrained areas. Across Bangladesh, the widespread use of mobile phones and the limited availability of traditional mental health resources offer the potential for mobile applications to be a scalable means of improving access to mental health care (Alam et al., 2025; Ahmed et al., 2020; Khatun et al., 2014; Khatun et al., 2015). Digital mental health (DMH) refers to the delivery of mental health education, self-management, screening, monitoring, and treatment (Torous et al., 2025; Fitzpatrick 2023; Chou et al., 2026) through digital platforms such as smartphone applications, on-line materials, telehealth, and related technologies. A systematic review and meta-analysis of randomized controlled trials in LMICs found moderate-to-large reductions in depression and anxiety symptoms as a result of digital mental health interventions (Kim et al., 2023). However, there is doubt as to its long-term efficacy and practical implementation issues in low-resource communities (Torous et al., 2025; Alagarajah et al., 2024). Therefore, it is critical to have an understanding of the predictors of adoption and continued usage of smartphone-based mental health applications to ensure their successful implementation in Bangladesh.

The ability to seek, understand, assess, and purposefully and effectively access digital mental health resources is becoming an important factor in technology use in mental health care (Del Pilar Arias López et al., 2023; Ban et al., 2024). This construct builds on mental health literacy as defined by Jorm et al., (1997), which refers to the ability to recognize mental disorders and understand the effectiveness of treatment, to encompass the skills required to navigate, evaluate, and employ digital tools. This is significant because knowing that there is a problem of depression does not necessarily mean that one can discern which apps are trustworthy or whether apps are evidence-based (Torous et al., 2022; Morton et al., 2021). Cross-national studies confirm that digital literacy can be a strong driver of mobile health app use, even more than privacy fears (Fan et al., 2023). Digital mental health literacy is recognized as a key extension to the TAM that could facilitate the elimination of barriers to the use of digital mental health (Adnan et al., 2025). In addition, digital mental health literacy is a modifiable construct, as targeting digital literacy can increase mental health literacy, decrease stigma, and increase help-seeking attitudes and mental health outcomes (Yeo et al., 2023; Chen et al., 2024). Lack of awareness of digital mental health tools has been detected as one of the major challenges that need to be addressed in Bangladesh, especially in urban slum area (Alam et al., 2025). On the other hand, digital health literacy typically correlates with greater access to digital health tools and services among populations and contexts (Cheng et al., 2022; Yuen et al., 2024; Shaw et al., 2024).

According to the Technology Acceptance Model (TAM), users’ willingness to use information technology is influenced more by two cognitive beliefs: perceived usefulness and perceived ease of use (Davis, 1989). In general, the larger direct relationship is expected to be between perceived usefulness and use intention to technology use, and a partial indirect relationship may be expected between perceived ease of use and use intention to technology use via its effect on perceived usefulness (Davis, 1989). This baseline pattern is also frequently found in mobile health and mental health systems; TAM has proven to be a robust model to explain user intent to use mental health apps, as found in several studies (Adnan et al., 2025). For instance, in Bangladesh, among university students, perceived usefulness and social influence were found to explain higher intention to adopt digital health for mental health, while perceived ease of use explained general intention to adopt digital health (Sifat et al., 2022). The results indicated that in Bangladesh, there is a cultural relevance of TAM constructs as predictors of the adoption of digital mental health.

The factors explaining mental health app usage are technological, individual, and social. Research also suggests that willingness is a close approximation to behavioral intention, which is similar to the TAM constructs (Walle et al., 2023; Sifat et al., 2022). Perceived usefulness is a consistent driver, where users preferred applications that enabled them to track their symptoms, were affordable, and had time-saving features over traditional applications (Alina & Zarlis, 2026). Global interest in mental health apps is high: in a recent meta-analysis, prospective use of mental health apps by the respondents was reported as 79%, with only 23.3% reporting current use, and a high percentage of 72.8% thought that mental health apps were useful (Guracho et al., 2023). This intention-behavior gap is significant, and the actual barriers are high awareness, inadequate onboarding, varying quality of the app, cost sensitivity, technical issues, and poor engagement after the download (Sawrikar & Mote, 2022; Chan & Honey, 2021; Szinay et al., 2020). The majority of users feel that mental health apps are complementary to rather than substitutes for traditional treatment services (Chan & Honey, 2021). Most importantly, for those who experience mental health stigma (a common concern in Bangladesh), smartphone applications could provide a level of privacy and accessibility that help to make seeking assistance easier (Kim et al., 2022).

Sociodemographic factors play a significant role in shaping inequalities in digital access, digital literacy, and eagerness to use mental health apps. The most consistent predictor is age – generally, older age corresponds to less adoption of apps and less digital health literacy (Tartaglia et al., 2024; Carroll et al., 2017; Ranjani et al., 2026); however, older adults who have frequent digital experiences tend to have greater digital health literacy than those who have limited digital experiences (Hoffman, 2026; Shi et al., 2024). Education is also consistently linked to digital health adoption, with higher levels of education being associated with increased app use, better acceptance, and increased digital literacy, likely due to greater digital skills and information processing capacity (Carroll et al., 2017; Fan et al., 2023; Schomakers et al., 2021). Findings on sex differences in mental health app use are inconsistent and are dependent on context; some studies indicate higher usage among females (Cruz et al., 2023; Sifat et al., 2022), and other studies in India found high willingness among males (Ranjani et al., 2026). Nevertheless, in Bangladesh, there is some evidence that women’s personal ownership of a smartphone and awareness of the technology may be lower than men’s, but that women are similarly or more likely to use an app to help them with their mental health when access is equated (Sifat et al., 2022; Tran et al., 2015). Digital opportunity and willingness are strongly shaped by urban/rural residence and income, as older adults, women, and rural residents have the lowest level of digital access across LMICs (Hui et al., 2022). In Ethiopia, mobile internet use and urban residence independently correlated with willingness to use mobile health applications (Walle et al., 2023). Likewise, in Bangladesh, the use and delivery of digital mental health services are severely hindered by the digital access and connectivity of rural and economically disadvantaged communities and their lack of access to organizations supporting them (Koly et al., 2022). Importantly, smartphone ownership is not synonymous with digital readiness: while 92% of households had access to a smartphone in a Dhaka urban slum, only 45% of the households had a smartphone personally owned (Alam et al., 2025).

Although digital mental health has seen increased interest in Bangladesh, evidence of this is limited, especially at the population level (Koly et al., 2022; Alam et al., 2024; Sifat et al., 2022). There are fewer studies that have explored digital mental health literacy, Technology Acceptance Model constructs, and sociodemographic characteristics within an integrated framework (Adnan et al., 2025; Torous et al., 2025). The evidence specific to willingness to use mental health apps on smartphones is limited (Guracho et al., 2023). This evidence gap is important as the prevalence of depression and anxiety is high in Bangladesh and mental health services are limited (Alam et al., 2024; Koly et al., 2022), and smartphone penetration is increasing. Therefore, it is important to understand the correlates of willingness to adopt digital mental health tools early on in designing implementation strategies and identifying the populations that might benefit most from digital interventions.

This study aimed to explore the factors influencing ready-to-use mental health applications by Bangladeshi adults in urban, semi-urban, and rural areas using smartphones. The main goal was to explore the relationships between mental health literacy, perceived usefulness, perceived ease of use, and willingness to use mental health applications on a smartphone. The secondary goal was to explore relationships between sociodemographic factors and the willingness to use such applications. We hypothesized that (1) higher willingness would be associated with higher digital mental health literacy, (2) higher perceived usefulness and perceived ease of use would be associated with higher digital mental health literacy willingness, and (3) willingness would vary significantly by sociodemographic characteristics such as age, education, sex, residence, and income (Gao et al., 2018; Sifat et al., 2022; Chou et al., 2026). This study combines digital mental health literacy, constructs from the Technology Acceptance Model, and sociodemographic factors to offer a more holistic picture of the willingness to use mental health apps in an LMIC setting. The results could guide the design, adaptation, and equitable utilization of a digital mental health service for Bangladesh and other resource-limited environments with similar epidemiological and technological characteristics (Koly et al., 2022; Torous et al., 2025; Chou et al., 2026).

## 2. Materials and Methods

### 2.1 Study Design and Setting

This was an analytical cross-sectional study, carried out with paper-based data collection, with different divisions in three different areas of Bangladesh: urban, semi-urban, and rural. The data were gathered during the period of 1st February to 20th June 2026, in the Government Upazila Health Complexes and Community Primary Care Clinics at Dhaka (urban), Chattogram (semi-urban), and Sylhet (rural) districts. All instruments were administered in one-on-one interview sessions in the Bangla language. Details of the study site selection, the recruitment of participants, sampling methodology, and baseline sample characteristics (n = 400) are reported in the companion paper (Part I) (Maruf et al., 2026)

### 2.2 Participants and Inclusion Criteria

Adults aged 18-60 years who attended primary care facilities were eligible to enroll if they had a current psychotic episode or severe drug abuse, were excluded, and were able to provide written informed consent in Bangla and use a smartphone regularly. Those who did not have a smartphone had their data analyzed in the companion paper but were not included in logistic regression models presented here.

### 2.3 Measurement Instruments

Patients were evaluated for depression and anxiety by Patient Health Questionnaire-9 (PHQ-9, 0–27) and Generalized Anxiety Disorder-7 (GAD-7, 0–21), respectively, which are Bangla-validated. The total scores from the baseline questionnaires (PHQ-9 and GAD-7) from the companion paper were not re-measured in the present analysis but were included as continuous independent variables.

Digital mental health literacy was evaluated by a 10-item scale designed by the researchers based on their measurements of awareness and knowledge of mental health apps, availability of online counselling, previous use of mental health apps, perceived effectiveness of mental health apps, and ability to download apps. Six items were scored (0 = no, 1 = yes), giving a range of 0 to 6 (with higher scores denoting higher levels of literacy; α = .762). The Technology Acceptance Model (TAM) scale, modified for mental health apps, included two subscales (Perceived Usefulness (PU) and Perceived Ease of Use (PEOU) each with 5 items and ranging from 5 to 25, with 1 = strongly disagree to 5 = strongly agree, α = .937 and α = .924, respectively.

The willingness to use mental health applications was measured by a 5-item behavioral intention scale adapted from UTAUT with a 5-point Likert scale (1-5; α = .946). Willingness was dichotomised at the mean score for primary analysis (≥3.5 = willing (1), <3.5 = not willing (0)).

### 2.4 Data Collection

A questionnaire was used for data collection, which included sections A (sociodemographic); B (PHQ-9); C (GAD-7); F (Digital access and digital literacy); G (TAM scale); H (Willingness to use mental health applications). The questionnaire was administered face-to-face by trained research assistants who interviewed the participants in Bangla for about 20–25 minutes.

### 2.5 Statistical Analysis

All scales and predictor variables were statistically described (Mean, Standard Deviation, Frequencies). All multi-item scales had internally consistent Cronbach’s alpha coefficients (all α ≥ .762). Spearman rank-order correlations were calculated between digital literacy, TAM subscales, depression, anxiety, and willingness; and chi-square tests were used to explore the relationship between categorical sociodemographic variables and willingness.

A hierarchical binary logistic regression model was fitted with willingness to use mental health apps (dichotomized) as the dependent variable. Independent variables were entered in a sequential fashion: Block 1, sociodemographic (age, sex, education, income, place of residence, marital status); Block 2, digital access and literacy score; Block 3, depression and anxiety severity score; Block 4, TAM sub-scores (perceived usefulness and perceived ease of use). Each block was reported with variance explained by the Nagelkerke R^2^ increments (ΔR^2^). All the variables were tested for multicollinearity using variance inflation factors (VIFs), with all values below 10. Adjusted odds ratios (aOR) with 95% confidence intervals (CI) and p-values are reported. Pre-specified stratified analysis by urban/rural residence and sex was performed. Data were analyzed using SPSS software version 27 (IBM Corp., Armonk, NY) with a two-tailed significance level of p < .05.

### 2.6 Ethics Approval

All participants were informed in Bangla and gave written informed consent before participating in this study, which received approval from Manabik Shahajya Sangstha (MSS), Ethical Review Committee (MSS-ERC), reference no. MSS-ERC-SFP-R-35/2026. The research followed the Declaration of Helsinki and the ethical principles of research on human subjects. Personally identifiable information was not collected in the analysis dataset, and all data were coded anonymously with individual participant codes. Paper questionnaires were placed in a locked cabinet that only authorized research personnel had access to.

## 3. Results

### 3.1 Sample Characteristics

Paper 2 analysis included all N = 400 participants from the combined dataset. All participants reported owning or regularly using a smartphone. The sample characteristics were identical to Paper 1, with a mean age of 40.22 years (SD = 10.14), nearly equal distribution by sex (33.2% male, 33.5% female, 33.2% missing), and equal stratification by education level (20% per category) and residence type (33.2% urban, 33.2% semi-urban, 33.5% rural).

### 3.2 Digital Literacy, Technology Acceptance, and Willingness Outcomes

Table 1 presents descriptive statistics for all Paper 2 outcome measures and predictors. The digital mental health literacy scores ranged from 0 to 6 (M = 5.36, SD = 2.77), suggesting moderately high mental health app and digital resource awareness. The mean score for the Perceived Usefulness subscale was very high (M = 17.55, SD = 4.40, range: 5–25), indicating a high perceived usefulness of mental health apps. Perceived Ease of Use also showed high scores (M = 17.55, SD = 4.42, range: 5–25). The willingness to use the mental health apps had a mean of 3.51 (SD = 0.88, range 1–5), with 235 participants (58.75%) indicating at or above a mean of 3.5, which corresponds with moderate to high willingness to use the app.

**Table 1.**
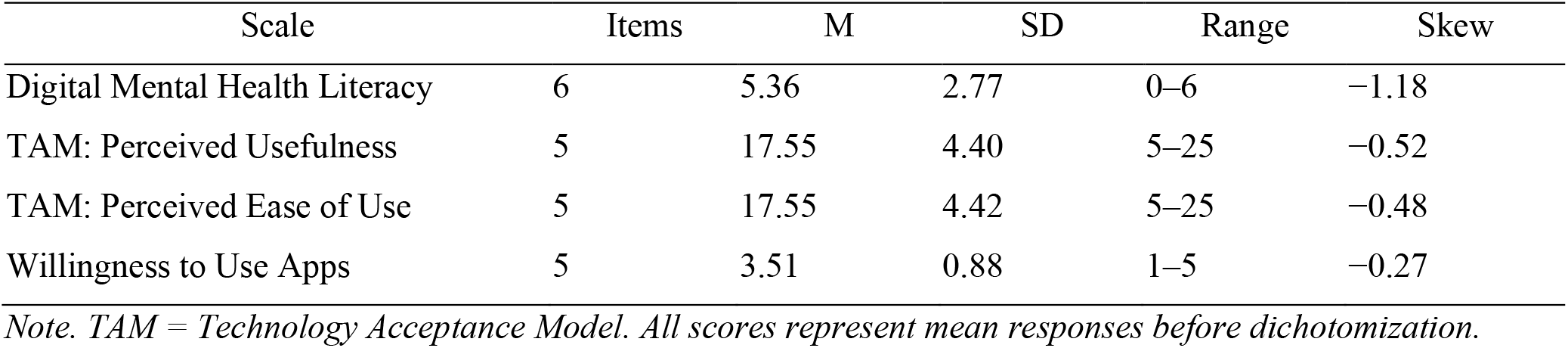
Descriptive Statistics for Paper 2 Scales: Digital Literacy, TAM, and Willingness (N = 400)

| Scale | Items | M | SD | Range | Skew |
| --- | --- | --- | --- | --- | --- |
| Digital Mental Health Literacy | 6 | 5.36 | 2.77 | 0–6 | –1.18 |
| TAM: Perceived Usefulness | 5 | 17.55 | 4.40 | 5–25 | –0.52 |
| TAM: Perceived Ease of Use | 5 | 17.55 | 4.42 | 5–25 | –0.48 |
| Willingness to Use Apps | 5 | 3.51 | 0.88 | 1–5 | –0.27 |

*Note.* TAM = Technology Acceptance Model. All scores represent mean responses before dichotomization.

### 3.3 Scale Reliability and Internal Consistency

Cronbach’s alpha coefficients were calculated for all multi-item scales. Table 2 presents internal consistency estimates. Good reliability was found for the digital mental health literacy scale (α = .762). The reliability of the TAM subscales was good: Perceived Usefulness (α = .937) and Perceived Ease of Use (α = .924). The Willingness-to-use scale had the highest internal consistency (α = .946). The depression scores on the PHQ-9 (α = .843) and the anxiety scores on the GAD-7 (α = .865) used to predict in the Paper 2 analysis also demonstrated good to excellent reliability.

**Table 2.**
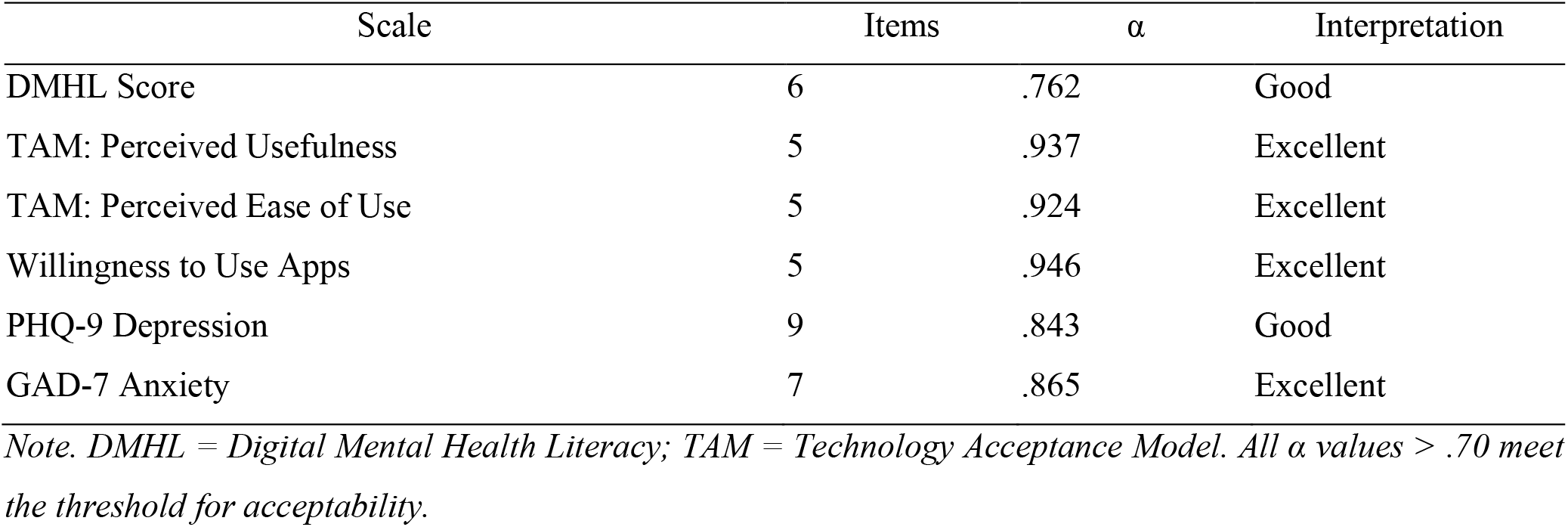
Reliability Coefficients (Cronbach’s Alpha) for Paper 2 Scales.

| Scale | Items | $\alpha$ | Interpretation |
| --- | --- | --- | --- |
| DMHL Score | 6 | .762 | Good |
| TAM: Perceived Usefulness | 5 | .937 | Excellent |
| TAM: Perceived Ease of Use | 5 | .924 | Excellent |
| Willingness to Use Apps | 5 | .946 | Excellent |
| PHQ-9 Depression | 9 | .843 | Good |
| GAD-7 Anxiety | 7 | .865 | Excellent |

*Note.* DMHL = Digital Mental Health Literacy; TAM = Technology Acceptance Model. All $\alpha$ values $> .70$ meet the threshold for acceptability.

### 3.4 Hierarchical Binary Logistic Regression Analysis

The complete hierarchical regression model is shown in Table 3, including model statistics and incremental variance explained. Willingness to use mental health apps was dichotomized using the mean score (≥ 3.5 = willing and < 3.5 = not willing) as the dependent variable in a hierarchical binary logistic regression. Four sets of independent variables were entered one after another: Block 1 (Sociodemographic: Age, Sex, Education, Employment, Residence, Marital Status); Block 2 (Digital access and literacy); Block 3 (Mental health status: PHQ-9 and GAD-7 score); Block 4 (TAM predictors: Perceived Usefulness and Perceived Ease of Use).

**Table 3.** Hierarchical Binary Logistic Regression: Predictors of Willingness to Use Mental Health Apps.

| Predictor | aOR | 95% CI | p | Sig. | $\Delta R^2$ |
| --- | --- | --- | --- | --- | --- |
| Block 1: Sociodemographics |  |  |  |  | .042 |
| Age (per year) | 1.02 | 0.99–1.05 | .218 | ns |  |
| Sex (Female vs. Male) | 1.09 | 0.62–1.92 | .754 | ns |  |
| Block 2: Digital Access & Literacy |  |  |  |  | .128 |
| DMHL Score (per point) | 1.24 | 1.11–1.39 | <.001 | *** |  |
| Block 3: Mental Health Status |  |  |  |  | .086 |
| PHQ-9 Depression Score | 0.98 | 0.94–1.02 | .363 | ns |  |
| GAD-7 Anxiety Score | 1.08 | 1.03–1.14 | .004 | ** |  |
| Block 4: TAM Predictors |  |  |  |  | .321 |
| TAM: Perceived Usefulness | 1.29 | 1.19–1.41 | <.001 | *** |  |
| TAM: Perceived Ease of Use | 1.17 | 1.08–1.27 | <.001 | *** |  |
| Full Model |  |  |  |  | .577 |

*Note.* aOR = adjusted odds ratio; CI = confidence interval; $\Delta R^2$ = Nagelkerke $R^2$ change from previous block; DMHL = Digital Mental Health Literacy; TAM = Technology Acceptance Model. \*\*\* $p < .001$ , \*\* $p < .01$ , ns = not significant. Full model $R^2$ (Nagelkerke) = .577.

### 3.5 Model Interpretation and Key Findings

The hierarchical logistic regression model explained 57.7% of the variance in willingness to use mental health apps (Nagelkerke R^2^ = .577). The explained variance was due to sociodemographic factors (Block 1) was 4.2%. The greatest added explained variance was for digital mental health literacy (12.8%, Block 2). Each additional point on the digital literacy scale increased odds of willingness by 24% (aOR = 1.24, 95% CI = 1.11–1.39, p < .001). Another 8.6% of variance was contributed by mental health status (Block 3). It is noteworthy that increased anxiety symptoms (GAD-7) were independently associated with increased willingness, with an aOR of 1.08 per point (95% CI = 1.03–1.14, p = .004), whereas depression symptoms were not independently associated. TAM predictors (Block 4) demonstrated the largest effect, explaining an additional 32.1% of variance beyond all other blocks. Perceived Usefulness (aOR = 1.29, 95% CI = 1.19–1.41, p < .001) and Perceived Ease of Use (aOR = 1.17, 95% CI = 1.08–1.27, p < .001) were both highly significant independent predictors, with each unit increase on either subscale substantially increasing willingness odds.

### 3.6 Summary of Findings and Implications

This analysis revealed several evidence-informed pathways of increased willingness to use mental health apps in Bangladesh. Digital mental health literacy is a key upstream predictor, and publicity about the availability, capabilities and benefits of mental health apps could increase uptake. Higher levels of anxiety symptoms – which may indicate a higher mental health need – were linked to greater willingness, which is interesting as it indicates that the more people were experiencing anxiety, the more they knew that digital mental health tools could be useful. Perhaps most importantly, the constructs of Perceived Usefulness and Perceived Ease of Use in the Technology Acceptance Model were most prominent in predicting willingness. The implications for practice are that user-friendly app design interfaces with a clear illustration of the benefits of mental health should be the focus of interventions to improve uptake and engagement.

## 4. Discussion

The current study aimed to explore factors related to the willingness to use the Apps for mental health among adults in Bangladesh involving the constructs of digital mental health literacy (DMHL), perceived usefulness and perceived ease of use of the Technology Acceptance Model (TAM), depression and anxiety severity, along with the soci-demographic factors. In general, DMHL, perceived usefulness and perceived ease of use each had a positive association with willingness to use mental health applications, independently. Willingness was also positively related to anxiety symptoms. After controlling for other factors, depression symptoms, age and sex were not independently related to willingness. These results indicate that the factors influencing the willingness to use digital mental health technologies are more closely linked to the participants’ digital capabilities and their confidence about the usefulness of the technologies than to demographic data.

### Digital Mental Health Literacy

The odds ratio for association between DMHL and willingness (1.24) is similar to that of high-income nations. The findings of Ochnik et al. (2024), regarding patient depression and anxiety acceptance of technology, were similar to the current study, with digital health literacy being a significant predictor, but with similar effect sizes, in Poland. Fan et al. (2023) conducted a cross-national study and found that privacy concerns are not as strong as digital literacy in predicting mHealth app adoption, which has a similar result as the present study with an emphasis on DMHL rather than demographic factors. There is limited evidence in Bangladesh, yet a study by Alam et al. (2025) found that the awareness of digital health tools was poor, especially in urban slums, and a study by Sifat et al. (2022) showed that a short educational intervention could be used to help a university student improve their knowledge of digital health. The present population-based observation adds to the earlier work, and helps justify the need to focus on DMHL-building interventions as a scalable approach to the adoption of these interventions.

### Technology Acceptance Model Constructs

As TAM suggested, both perceived usefulness (aOR = 1.29) and perceived ease of use (aOR = 1.17) were independently associated with willingness. This is different from some mobile health literature that found that PEOU had weaker or indirect relationships (Mahajan et al., 2025); and in the current study, both constructs were related to willingness independently, indicating users in Bangladesh consider both utility and usability in mental health apps. These patterns were also found among Bangladeshi university students, where perceived usefulness positively predicted the intention to use digital health for mental health (β = 0.43), and perceived ease of use predicted general digital health user intentions but not for mental health-specific use (Sifat et al., 2022). The present study also shows that both TAM constructs predict, even in a population-based sample with lower-literacy and rural respondents which is a factor that makes it unique and furthering the argument that the constructs are generalizable beyond the educated urban population. But the significant intention-behavior gap reported internationally has grave implications: Guracho et al. (2023) analysed 21 previous studies and identified a 79– 23.3% gap between intention to use mental health apps and actual use. This gap indicates that while willingness is important, it is not enough if it is not accompanied by overcoming post-adoption issues like quality of the application, personalization, clinical integration, and support for continued engagement.

### Mental Health Symptoms and Willingness

Symptoms of anxiety were positively associated with willingness (aOR = 1.08, p < .05) but depression symptoms were not independently associated after adjustment. This differential pattern corroborates the international findings of mixed associations between the severity of symptoms and the adoption of digital health. In the study of qualitative interviews, Cruz et al. (2023) identified that privacy and accessibility were common reasons why people used mental health apps, while depressed people were less motivated and engaged in the apps. Kim et al. (2022) reported higher levels of endorsement of mental health apps as stigma reducing for those with anxiety than for those with depression. Koly et al. (2022) cited in the context of Bangladesh, that anxiety might make people choose online and asynchronously over in-person services because of stigma issues. This non-associability may be due to depression being a syndrome of behavioral withdrawal or decreased initiative that is not measured by willingness. To better determine the relationship between willingness and continued app use, longitudinal research designs using actual app engagement data are required.

### Sociodemographic Factors

In contrast to mental Health research primarily focused on demographics that suggest age and sex should predict, but not after accounting for DMHL constructs, and similar to more modern technology adoption models, age and sex did not independently predict willingness. Carroll et al., 2017, identified age as a significant predictor of app adoption in a large sample in the United States but did not control for perceptions about the app. When Schomakers et al., 2021, controlled for perceptions about the app, the effect of age substantially dampened. Sifat et al. (2022) found evidence of a difference in willingness, with women being more willing, in Bangladesh, but again, this study did not control for digital literacy and technology perceptions. The results of the present study indicate that there are two implications. First, demographics become insignificant once there are DMHL and positive perceptions about technology present; and second, interventions focusing on DMHL and perceptions of the technology’s design can be more effective than demographic targeting in enhancing adoption. This has equity implications, as enhancing digital literacy and app quality could bring technology to older citizens and rural women at large in a way that could help to level their access to the technology—an important consideration in Bangladesh, where rural women and older adults have disparities in digital access (Ahmed et al., 2020; Tran et al., 2015).

### Limitations

There are a number of significant limitations in this research. First, the cross-sectional design eliminates drawing causal inferences on the relationship between DMHL, technology acceptance and mental health symptoms and willingness to use applications. Second, willingness was measured as behavioral intentions not sustained or actual use; willingness does not necessarily mean sustained use or engagement in real-world use. Third, self-reported measures can have social-desirability bias, especially for measures of mental health. Fourth, willingness was dichotomized at the mean score (willingness ≥3.5 and willingness < 3.5), potentially diminishing the information on the original continuous scale. Five, the study did not evaluate structural barriers to implementation such as affordability, the reliability of internet access, data privacy issues, language availability, smartphone storage space, or integration with the health care system, which may result in significant constraints to actual implementation even if uptake is high. Sixth, the study quantified the awareness and knowledge of digital mental health resources, but did not measure actual use or retention of resources, which should be measured through app analytics and through longitudinal follow-up. Lastly, the results are only applicable to the three divisions of Bangladesh and cannot be extrapolated to the rural areas of Bangladesh, or to resource poor areas of the world, without similar digital infrastructure.

### Implications for Intervention

The results have significant implications for the implementation of digital mental health services in Bangladesh. Public health promotion strategies and initiatives to increase digital mental health literacy should be prioritized by addressing the needs of adults who are using mental health apps and online counselling, including campaigns that reach low-literacy and rural populations. Meta-analyses indicate that short (15-30 min) community health worker-led educational sessions are effective at improving digital literacy and decreasing stigma (Yeo et al. 2023; Chen, et al., 2024). Second, app developers must make the features perceived as useful to the users – by providing symptom tracking, affordability, evidence-based content, Bangla language interface, and integration with primary care (Sawrikar & Mote, 2022). Third, user-centered design and iterative testing at different levels of literacy is needed; 40% of users stop using apps after one week because they didn’t know how to use them (Szinay et al., 2020). Fourth, engagement features (such as reminders, progress visualization, and clinician endorsement) that have been shown to increase retention (Chan & Honey, 2021) should be used with early adopters (anxiety patients, high DMHL individuals) in implementation pilots. Fifth, interventions need to measure actual adoption and retention using app analytics and longitudinal designs, because the intention-behaviour gap is largely due to app quality and clinical integration rather than unwillingness (Walle et al., 2023). Last, structural barriers need to be addressed: subsidization of access by low-income users, reliability of mobile internet, protecting privacy, and integration with existing upazila health complexes and community clinics (Hui et al., 2022). If it is recognized as a condition that is modifiable, and individual and structural factors are systematically tackled, then Bangladesh will be able to scale up evidence-based mental health services to the underserved populations.

## 5. Conclusion

In this study, digital mental health literacy, perceived usefulness, and perceived ease of use were found to be independent predictors of willingness to use smartphone-based mental health applications among Bangladeshi adult population. Importantly, demographic characteristics (age, sex) were not independently predictive after adjustment for constructs of DMHL; this has important equity implications. Targeted interventions to enhance digital literacy and app quality can level the playing field when it comes to uptake, rather than demographics being a barrier. This significant discrepancy between intention and actual use has been reported in previous studies and indicates that post-adoption challenges, such as app quality, clinical integration, data reliability, protections, and user engagement design, must be addressed systematically. Although consumers are willing, the ICT infrastructure in Bangladesh is still weak and hinders implementation efforts, which will need cooperation among the public health, telecom, app development and healthcare communities. While there is a limited body of research on digital mental health adoption in South Asia, this study shows that TAM constructs are still valid predictors in diverse population-based settings with limited resources. Digital mental health literacy introduces a new dimension to the frameworks of technology acceptance and identifies a target for intervention that can be modified. However, there is a need for longitudinal studies using real app usage data, implementation science studies and embedded effectiveness trials to better understand the causality and maximize actual outcomes in the real world. While digital mental health is a scalable approach to fill Bangladesh’s mental health treatment gap, its implementation should target both individual and structural barriers – such as perceptions of technology and literacy, and access and infrastructure, as well as clinical integration. Future research and policy need to focus on developing digital capabilities, alongside improving the quality of the apps and readiness of health systems.

## Data Availability

All data produced in the present study are available upon reasonable request to the authors

## Acknowledgments

We acknowledge the support of government upazila health complexes and community primary care clinics in Dhaka, Chattogram, and Sylhet divisions for facilitating data collection. We thank the research assistants who conducted participant interviews and data entry. We acknowledge the Bangladesh Medical Research Council for ethical oversight.

## Declaration of AI Use

The authors have also, as part of the preparation of this work, resorted to the help of Claude, an artificial intelligence language model, for writing and as an organizational assistant. The responsibility for the scientific content, data analysis, interpretation and conclusions of the work remain unchanged and lie with the human authors, who have the authority to decide on the results of these analyses, interpretations and conclusions. All AI-assisted text is thoroughly reviewed by the authors and the accuracy and integrity of this manuscript are their sole responsibility.

## Author Contributions

Sabbir Maruf: Conceptualization, Study Design, Methodology, Data Collection, Supervision, Data Analysis, Writing - Original Draft, Writing - Review & Editing. Habib: Conceptualization, Methodology, Writing - Review & Editing. Faria: Data Collection, Introduction and Conclusion. Shakila: Results and Discussion. Mamunur: Abstract and Conclusion; also, Masum and Bayezid: Data Collection

## Data Availability Statement

The datasets generated during the current study are not publicly available due to participant confidentiality and ethical restrictions but are available from the corresponding author on reasonable request with appropriate institutional ethics approval and data sharing agreements.

